# Persistent symptoms among children and adolescents with and without anti-SARS-CoV-2 antibodies: a population-based serological study in Geneva, Switzerland

**DOI:** 10.1101/2021.12.23.21268298

**Authors:** Roxane Dumont, Mayssam Nehme, Elsa Lorthe, Carlos de Mestral, Viviane Richard, Hélène Baysson, Francesco Pennacchio, Julien Lamour, Claire Semaani, María-Eugenia Zaballa, Nick Pullen, Anne Perrin, Arnaud G. L’Huillier, Klara M. Posfay-Barbe, Idris Guessous, Silvia Stringhini, on behalf of the Specchio-COVID19 study group

## Abstract

**Background:** It is now established that a significant proportion of adults experience persistent symptoms after SARS-CoV-2 infection. However, evidence for children and adolescents is still inconclusive. In this population-based study, we examine the proportion of children and adolescents reporting persistent symptoms after SARS-CoV-2 infection, as assessed by serological status, and compare this to a seronegative control group.

**Methods:** We conducted a serosurvey in June-July 2021, recruiting 660 children and adolescents from 391 households selected randomly from the Geneva population. We tested participants for anti-SARS-CoV-2 antibodies targeting the nucleocapsid (N) protein to determine previous infection. A parent filled a questionnaire including questions on COVID-19-related symptoms lasting at least 2 weeks.

**Findings:** Among children seropositive for anti-SARS-CoV-2 antibodies, the sex- and age-adjusted prevalence of symptoms lasting longer than two weeks was 18.3%, compared to 11.1% among seronegative children (prevalence difference (ΔaPrev)=7.2%, 95%CI:1.5-13.0). Main symptoms declared among seropositive children were fatigue (11.5%) and headache (11.1%). For 8.6% (aPrev, 95%CI: 4.7-12.5) of seropositives, these symptoms were declared to be highly limiting of daily activities. Adolescents aged 12-17 years had a higher adjusted prevalence of persistent symptoms (aPrev=29.1%, 95%CI:19.4-38.7) than younger children. Comparing seropositive and seronegative adolescents, the estimated prevalence of symptoms lasting over four weeks is 4.4% (ΔaPrev, 95%CI:-3.8-13.6).

**Interpretation:** A significant proportion of children aged 12 to 17 years had symptoms lasting over two weeks after SARS-CoV-2 infection, with an estimated prevalence of symptoms lasting over 4 weeks of 4.4% in this age group. This represents a large number of adolescents in absolute terms, and should raise concern in the context of unknown long-term evolution of symptoms. Younger children appear to experience long-lasting symptoms less frequently, as no difference was observed between the seropositive and seronegative sample. Further studies with larger samples sizes are needed.

**Funding:** Swiss Federal Office of Public Health, Geneva General Directorate of Health, HUG Private Foundation, SSPH+, Fondation des Grangettes.

## Introduction

Long COVID or post-COVID syndrome refers to individuals who experience long-lasting symptoms weeks to months following a SARS-CoV-2 infection [1-2]. Evidence to date indicates that more than one third of adults who had mild to severe COVID-19 experience persistent symptoms several months after infection [3–6]. Risk factors include female sex, middle age, comorbidities and the number of symptoms in the acute phase [7]. However, young and previously healthy persons are also frequently affected [8]. As such, long COVID represents an increasing public health concern, potentially preventing affected individuals from returning to the workforce and continuing to burden the healthcare system. If substantial evidence is starting to emerge around long COVID in adults, pediatric long COVID has received much less attention [1]-[9]. To date, there is no generalized definition, nor a recognized diagnostic test for long COVID among children and no precise understanding of the duration of symptoms that justifies the diagnosis.

The pandemic has profoundly impacted directly or indirectly the lives of children and adolescents worldwide in terms of daily life and habits, mental and physical health, social behaviors and schooling [10]. Although they appear to be less susceptible to severe forms of COVID-19 compared to adults [11], recent evidence suggests that an undetermined proportion of infected children may also experience persistent symptoms post infection. Studies from Australia [12], Italy [13], England and Wales [3], and Switzerland [14] have reported prevalence estimates of persistent symptoms lasting more than 4 weeks ranging from 4.6% to 24.0%. The most frequently reported symptoms are fatigue, insomnia, respiratory symptoms (including chest pain and tightness), nasal congestion, muscle and joint pain and difficulty concentrating, which have been reported to last from four weeks up to 4 months [3], [13-15]. These studies differed considerably in their methodological approaches, their assessment of previous infection, sample size, and follow-up periods, so that the actual prevalence of long-lasting symptoms among children and adolescents is still debated. Importantly, most studies were based on clinical samples of confirmed cases, while only one study [14] included data from a population-based serosurvey. Population-based serological studies have the advantage of including, by definition, a representative sample of the population which includes severe, mild and asymptomatic infections, giving a more accurate estimation of the number of children infected (denominator). The difference between seropositive and seronegative also allows the distinction of persistent symptoms due to COVID-19 from symptoms due to other viruses. Understanding the frequency, duration and severity of persistent symptoms following SARS-CoV-2 infection in children is essential to guide public health strategies targeting this age group (for example, preventive measures in schools or vaccination programs), as well as to monitor the long-term physical and mental health impact of the pandemic. Using a representative sample of the general population of the canton of Geneva, Switzerland, we aimed to determine the proportion of children and adolescents reporting persistent symptoms after SARS-CoV-2 infection, as assessed by serological status, compared to seronegative children and adolescents and to identify risk factors of experiencing persistent symptoms.

## Methods

### Study population

Participants from a random sample of the general population of the state of Geneva, Switzerland, were invited to take part in a serological survey between June 1^st^ and July 7^th^, 2021 [16]. Lists of residents were provided by the Swiss Federal Office of Statistics within the framework of the Corona Immunitas Research project [17]. As per the study protocol, randomly selected children and adolescents aged between 6 months and 17 years were invited to participate with their family members. Overall, 22.3% of the invited families participated (Figure S1). After providing written informed consent, participants provided a venous blood sample. In each family, one of the parents completed a socio-demographic online questionnaire about themselves, the household and their child(ren). For children from 2 years old, the questionnaire included questions on COVID-19 persistent symptoms.

### Measures

Parents were asked if their child(ren) had experienced symptoms lasting at least 2 weeks. Parents could choose from an exhaustive list of 37 symptoms based on the literature at the time, grouped into six general categories of symptoms: fatigue, respiratory, gastrointestinal, musculoskeletal, neurological and dermatological symptoms (Table S1). For each symptom, details on the duration (2 to 3 weeks, 3 to 4 weeks, or more than 4 weeks) and impact on the child’s daily activities were collected. The impact/limitation of each selected symptom was based on the following question: “On a scale of 1 to 10, to what extent is this symptom limiting the child’s usual activities (attendance at school, nursery, studies, sports, games, etc.)? (1 very weak limitation - 10 strong limitation)”. We subsequently created a dichotomous variable where symptoms rated 6 or higher were considered as having a strong limitation on the child’s daily life.

Serological tests were based on the semiquantitative commercially-available immunoassay Roche Elecsys anti-SARS-CoV-2 N and seropositivity was defined using the manufacturer’s provided cut-off index of ≥1.0 [18] (See supplementary material). The Roche N serological test identifies anti-N antibodies, which are not produced following vaccination with mRNA vaccines used in Geneva to date, such as the Pfizer-BioNTech BNT162b2 and Moderna mRNA 1273 vaccines, and are therefore only detected if the individual was infected with SARS-CoV-2. Using this test allowed us to remove the effect of vaccination.

Parental education was categorized as primary for compulsory schooling, secondary for apprenticeship and high school, and tertiary for university. Household financial status was defined as average to poor if participants chose one of the following statements about their financial situation: “I have to be careful with my expenses and an unexpected event could put me into financial difficulty” or “I cannot cover my needs with my income and I need external support”.

### Statistical analysis

We compared the distribution of sociodemographic and COVID-19-related characteristics between children who tested positive for anti-SARS-CoV-2 antibodies and children who tested negative, overall and stratified into three age groups (2 to 5, 6 to 11 and 12 to 17). Participants with missing data were excluded. To estimate prevalence (95% confidence intervals) and prevalence differences, we used marginal prediction after logistic regression, adjusting for age and sex. To calculate prevalence ratios (95% CI), we used Poisson regression with robust variance, based on the sandwich estimator [19]. To account for the fact that many children were siblings, we conducted additional analyses on the prevalence ratio estimation using mixed-effect Poisson regression with robust variance [20]. Statistical significance was defined at a level of confidence of 95% and all analyses were performed with R (version 4.0.3).

### Results

Our sample comprised 660 children aged 2 to 17 years (49.4% girls, mean age 9.3 years [SD=4.5]) from 391 households (Table 1). A majority of parents (58.6%) had a tertiary education level, 7.6% had a primary education level, and 22.1% reported having an average to poor financial situation.

**Table 1.**
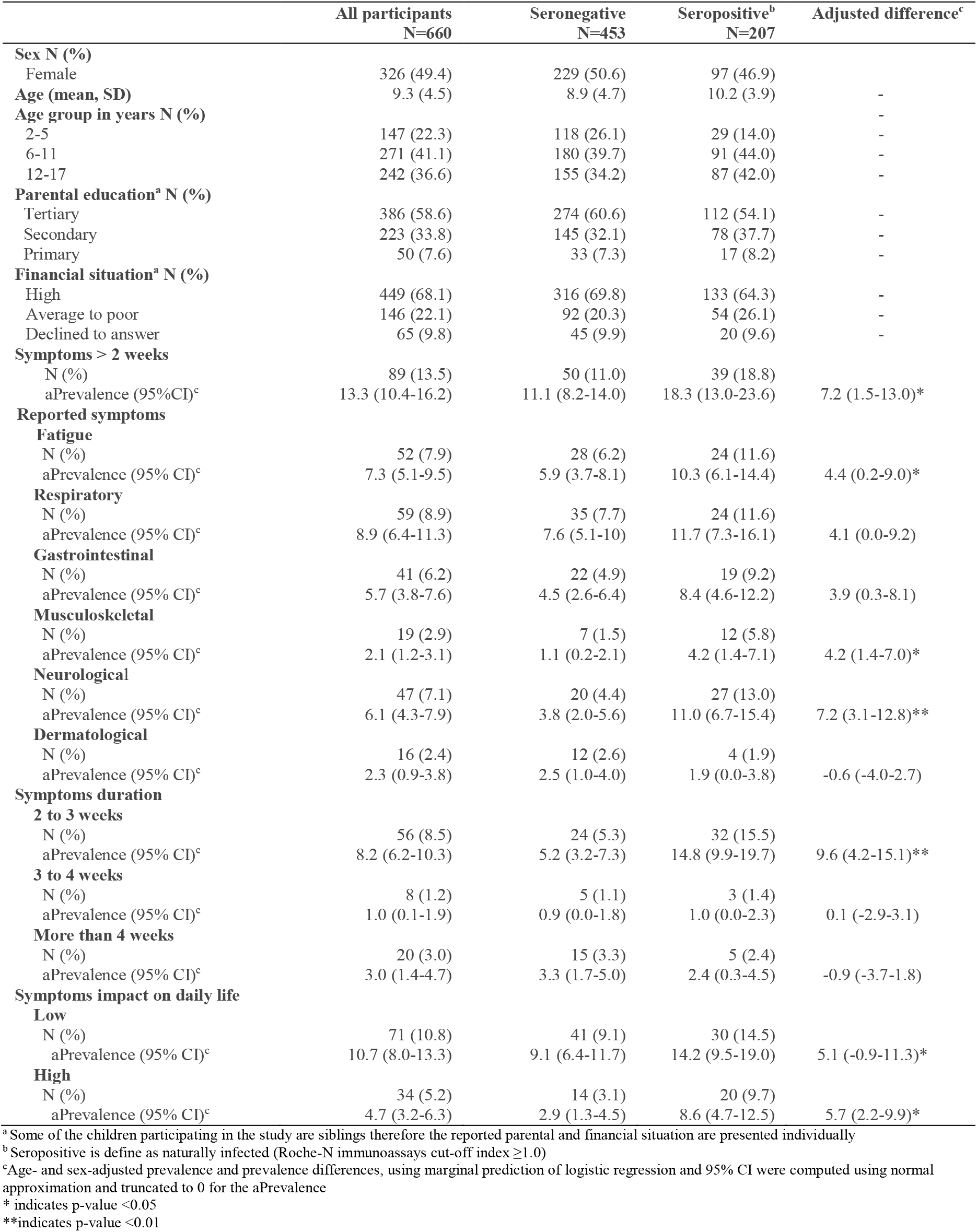
Descriptive statistics of the study population, stratified by serological status.

|  | All participants<br>N=660 | Seronegative<br>N=453 | Seropositive<sup>b</sup><br>N=207 | Adjusted difference<sup>c</sup> |
| --- | --- | --- | --- | --- |
| **Sex N (%)** |  |  |  |  |
| Female | 326 (49.4) | 229 (50.6) | 97 (46.9) |  |
| **Age (mean, SD)** | 9.3 (4.5) | 8.9 (4.7) | 10.2 (3.9) | - |
| **Age group in years N (%)** |  |  |  |  |
| 2-5 | 147 (22.3) | 118 (26.1) | 29 (14.0) | - |
| 6-11 | 271 (41.1) | 180 (39.7) | 91 (44.0) | - |
| 12-17 | 242 (36.6) | 155 (34.2) | 87 (42.0) | - |
| **Parental education<sup>a</sup> N (%)** |  |  |  |  |
| Tertiary | 386 (58.6) | 274 (60.6) | 112 (54.1) | - |
| Secondary | 223 (33.8) | 145 (32.1) | 78 (37.7) | - |
| Primary | 50 (7.6) | 33 (7.3) | 17 (8.2) | - |
| **Financial situation<sup>a</sup> N (%)** |  |  |  |  |
| High | 449 (68.1) | 316 (69.8) | 133 (64.3) | - |
| Average to poor | 146 (22.1) | 92 (20.3) | 54 (26.1) | - |
| Declined to answer | 65 (9.8) | 45 (9.9) | 20 (9.6) | - |
| **Symptoms > 2 weeks** |  |  |  |  |
| N (%) | 89 (13.5) | 50 (11.0) | 39 (18.8) |  |
| aPrevalence (95% CI)<sup>c</sup> | 13.3 (10.4-16.2) | 11.1 (8.2-14.0) | 18.3 (13.0-23.6) | 7.2 (1.5-13.0)* |
| **Reported symptoms** |  |  |  |  |
| **Fatigue** |  |  |  |  |
| N (%) | 52 (7.9) | 28 (6.2) | 24 (11.6) |  |
| aPrevalence (95% CI)<sup>c</sup> | 7.3 (5.1-9.5) | 5.9 (3.7-8.1) | 10.3 (6.1-14.4) | 4.4 (0.2-9.0)* |
| **Respiratory** |  |  |  |  |
| N (%) | 59 (8.9) | 35 (7.7) | 24 (11.6) |  |
| aPrevalence (95% CI)<sup>c</sup> | 8.9 (6.4-11.3) | 7.6 (5.1-10) | 11.7 (7.3-16.1) | 4.1 (0.0-9.2) |
| **Gastrointestinal** |  |  |  |  |
| N (%) | 41 (6.2) | 22 (4.9) | 19 (9.2) |  |
| aPrevalence (95% CI)<sup>c</sup> | 5.7 (3.8-7.6) | 4.5 (2.6-6.4) | 8.4 (4.6-12.2) | 3.9 (0.3-8.1) |
| **Musculoskeletal** |  |  |  |  |
| N (%) | 19 (2.9) | 7 (1.5) | 12 (5.8) |  |
| aPrevalence (95% CI)<sup>c</sup> | 2.1 (1.2-3.1) | 1.1 (0.2-2.1) | 4.2 (1.4-7.1) | 4.2 (1.4-7.0)* |
| **Neurological** |  |  |  |  |
| N (%) | 47 (7.1) | 20 (4.4) | 27 (13.0) |  |
| aPrevalence (95% CI)<sup>c</sup> | 6.1 (4.3-7.9) | 3.8 (2.0-5.6) | 11.0 (6.7-15.4) | 7.2 (3.1-12.8)** |
| **Dermatological** |  |  |  |  |
| N (%) | 16 (2.4) | 12 (2.6) | 4 (1.9) |  |
| aPrevalence (95% CI)<sup>c</sup> | 2.3 (0.9-3.8) | 2.5 (1.0-4.0) | 1.9 (0.0-3.8) | -0.6 (-4.0-2.7) |
| **Symptoms duration** |  |  |  |  |
| **2 to 3 weeks** |  |  |  |  |
| N (%) | 56 (8.5) | 24 (5.3) | 32 (15.5) |  |
| aPrevalence (95% CI)<sup>c</sup> | 8.2 (6.2-10.3) | 5.2 (3.2-7.3) | 14.8 (9.9-19.7) | 9.6 (4.2-15.1)** |
| **3 to 4 weeks** |  |  |  |  |
| N (%) | 8 (1.2) | 5 (1.1) | 3 (1.4) |  |
| aPrevalence (95% CI)<sup>c</sup> | 1.0 (0.1-1.9) | 0.9 (0.0-1.8) | 1.0 (0.0-2.3) | 0.1 (-2.9-3.1) |
| **More than 4 weeks** |  |  |  |  |
| N (%) | 20 (3.0) | 15 (3.3) | 5 (2.4) |  |
| aPrevalence (95% CI)<sup>c</sup> | 3.0 (1.4-4.7) | 3.3 (1.7-5.0) | 2.4 (0.3-4.5) | -0.9 (-3.7-1.8) |
| **Symptoms impact on daily life** |  |  |  |  |
| **Low** |  |  |  |  |
| N (%) | 71 (10.8) | 41 (9.1) | 30 (14.5) |  |
| aPrevalence (95% CI)<sup>c</sup> | 10.7 (8.0-13.3) | 9.1 (6.4-11.7) | 14.2 (9.5-19.0) | 5.1 (-0.9-11.3)* |
| **High** |  |  |  |  |
| N (%) | 34 (5.2) | 14 (3.1) | 20 (9.7) |  |
| aPrevalence (95% CI)<sup>c</sup> | 4.7 (3.2-6.3) | 2.9 (1.3-4.5) | 8.6 (4.7-12.5) | 5.7 (2.2-9.9)* |

<sup>a</sup> Some of the children participating in the study are siblings therefore the reported parental and financial situation are presented individually
<sup>b</sup> Seropositive is define as naturally infected (Roche-N immunoassays cut-off index $\geq 1.0$ )
<sup>c</sup> Age- and sex-adjusted prevalence and prevalence differences, using marginal prediction of logistic regression and 95% CI were computed using normal approximation and truncated to 0 for the aPrevalence
\* indicates p-value <0.05
\*\*indicates p-value <0.01

**Table 2.** Association between persistent symptoms and serological status.

| Report of symptoms lasting at least 2 weeks |  | No (%)<br>N=571 | Yes (%)<br>N=89 | Prevalence ratio<sup>a</sup><br>95% CI | Prevalence ratio<sup>b</sup><br>95% CI |
| --- | --- | --- | --- | --- | --- |
| **Sex** | Female | 285 (87.2) | 42 (12.8) | 1.0 (ref) | 1.0 (ref) |
|  | Male | 280 (85.6) | 47 (14.4) | 1.11 (0.76-1.64) | 1.14 (0.74-1.76) |
|  | Diverse | 8 (100.0) | 0 (0.0) | undefined | undefined |
| **Age (years)** | 2-5 | 132 (89.8) | 15 (10.2) | 1.0 (ref) | 1.0 (ref) |
|  | 6-11 | 236 (87.1) | 35 (12.9) | 1.26 (0.71-2.23) | 1.54 (0.30-7.83) |
|  | 12-17 | 203 (83.9) | 39 (16.1) | 1.57 (0.90-2.76) | 1.61 (0.86-2.98) |
| **Serological status** | Negative | 404 (89.0) | 50 (11.0) | 1.0 (ref) | 1.0 (ref) |
|  | Positive | 169 (81.2) | 39 (18.8) | 1.65 (1.13-2.41)*<sup>c</sup> | 1.62 (1.10-2.39)*<sup>c</sup> |

<sup>a</sup>Prevalence ratio and 95% confidence interval are from Poisson regression with robust variance, and are adjusted for age, sex, or both according to independent variable.
<sup>b</sup>Prevalence ratio and 95% confidence interval are from Poisson regression with robust variance and random effect on the household using the GLMMadaptive package in R, and are adjusted for age, sex, or both according to independent variable.
<sup>c</sup>Adjusting for age and sex
\* indicates p-value <0.05
\*\*indicates p-value <0.01

Overall, 13.5% of children (adjusted prevalence=13.3%, 95%CI: 10.4-16.2) were reported by their parent as having experienced at least one symptom that lasted more than two weeks since the beginning of the pandemic. Specifically, 18.3% (95%CI: 13.0-23.6) of seropositive children had symptoms lasting more than two weeks vs 11.1% (95%CI: 8.2-14.0), among seronegative children, with an adjusted prevalence difference of 7.2% (95%CI: 1.5-13.0) (Table 1). Among seropositive participants, 14.8% (95%CI: 9.9-19.7) reported symptoms lasting 2-3 weeks, 1.0% (95%CI: 0.0-2.3) 3-4 weeks and 2.4% (95%CI: 0.3-4.5) more than four weeks. Among seronegative participants, the corresponding adjusted prevalence were 5.2% (95%CI 3.2-7.3), 0.9% (95%CI: 0.0-1.8), and 3.3% (95%CI: 1.7-5.0). Moreover, for 8.6% (95%CI: 4.7-12.5) of seropositive children, these persistent symptoms were reported as highly limiting and resulting in a daily burden, compared to 2.9% (95%CI: 1.3-4.5) among seronegatives.

The most frequently reported symptoms by seropositive participants were fatigue (11.6%), headache (11.1%), fever (6.2%), runny nose (6.2%), loss of smell (4.8%) and loss of taste (3.8%). Among seronegatives, the most frequently reported symptoms were fatigue (6.2%), runny nose (5.9%), cough (4.2%) and sore throat (3.1%) (Table S1).

The age- and sex-adjusted prevalence of persistent symptoms varied across age groups (Figure 1, Table S2) and differed by serological status. Among seropositive adolescents aged 12 to 17 years, 29.0% (95%CI: 19.4-38.7) reported persistent symptoms, while the age-and-sex adjusted prevalence was 8.9% (95%CI: 4.4-13.4) among seronegative participants of the same age (Prevalence difference (ΔaPrev)=20.1%, 95%CI: 10.6-29.7). Differences between seropositive and seronegative participants were not significant among children aged 6 to 11 years (ΔaPrev =-0.5%, 95%CI: -8.2-7.1) and those aged 2 to 5 years (ΔaPrev =-0.8%, 95%CI: -13.4-11.6).

**Figure 1:**
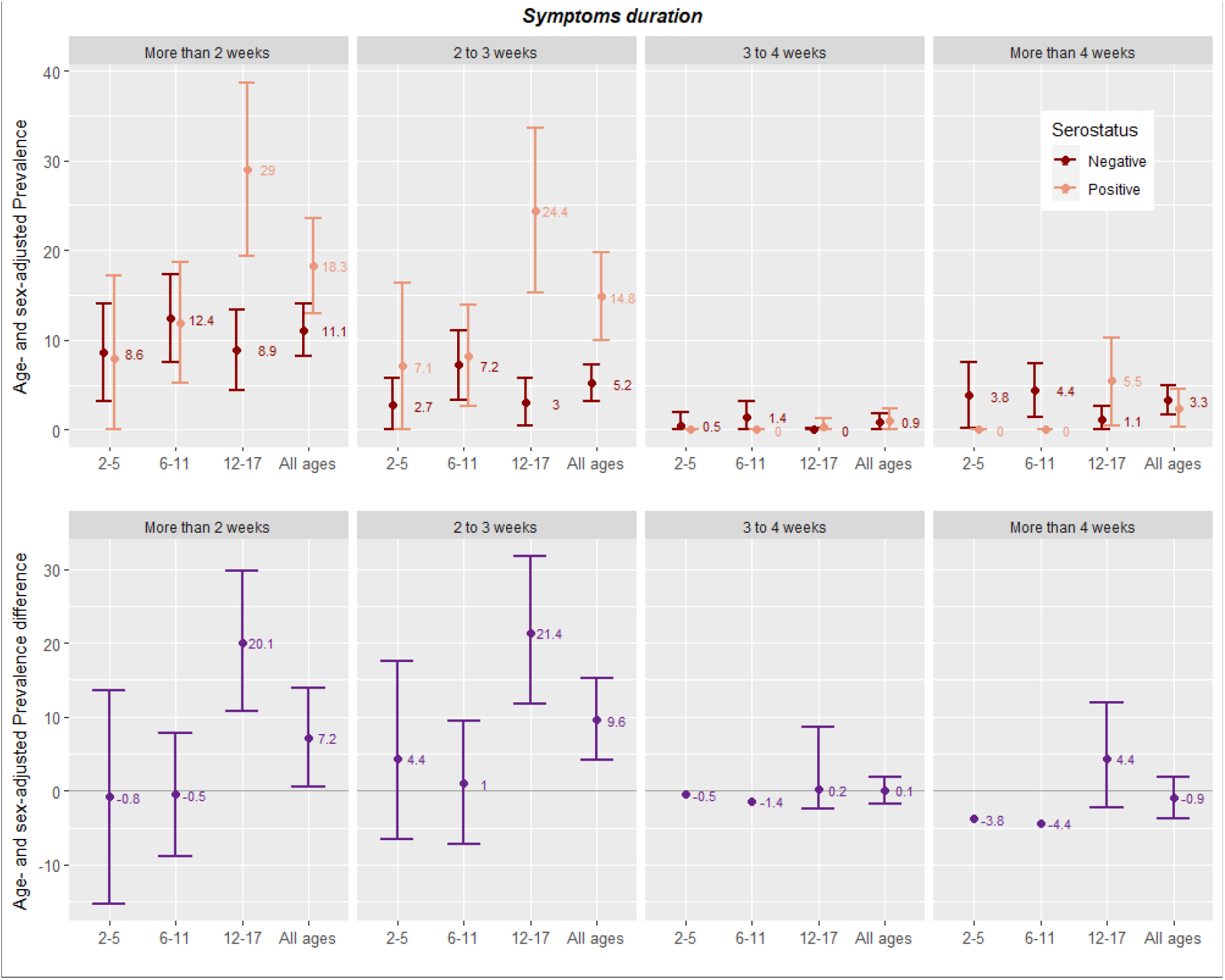
Age- and sex-adjusted prevalence and prevalence difference of persistent symptoms duration, stratified by age group and serological status.

Among seropositive adolescents aged 12 to 17 years, 5.5% (95%CI: 0.5-10.3) reported experiencing symptoms lasting longer than four weeks, while the prevalence among seronegative was 1.1% (95%CI: 0.0-2.6) with an adjusted prevalence difference in this age group of 4.4% (95%CI:-3.8-13.6). The most frequently reported symptoms category in this age group were fatigue (ΔaPrev =13.4%, 95%CI: 4.7-22.1), neurological (mostly headaches) (ΔaPrev =15.8%, 95%CI: 5.6-26.7) and respiratory symptoms (ΔaPrev =11.8%, 95%CI: 2.7-21.6), (Figure S2).

Overall, seropositive children and adolescents were 62% more likely than seronegatives to experience symptoms lasting more than 2 weeks (aPR = 1.62, 95%CI: 1.10-2.39). In sub-analyses focusing only on seropositive children, we observed that seropositive children whose parents had a primary education level were almost three times more likely (aPR = 2.97, 95%CI: 1.11-7.96) and those whose parents have a secondary education level twice as likely (aPR = 2.10, 95%CI: 1.07-4.12) to experience symptoms lasting more than two weeks, compared with seropositive children whose parents had a tertiary education level. Similarly, parents from households with an average to poor financial status were twice as likely to declare long-lasting symptoms for their children than parents with high financial status (aPR=2.18, 95%CI 1.09-4.35) (Table 3).

**Table 3.** Association between persistent symptoms and socioeconomic indicators among seropositive children.

| Report of symptoms lasting at least 2 weeks, among seropositive |  | No (%)<br>N=168 | Yes (%)<br>N=39 | Prevalence ratio<sup>a</sup><br>(95% CI) | Prevalence ratio<sup>b</sup><br>(95% CI) |
| --- | --- | --- | --- | --- | --- |
| **Sex** | Female | 80 (82.5) | 17 (17.5) | 1.0 (ref) | 1.0 (ref) |
|  | Male | 85 (79.4) | 22 (20.6) | 1.17 (0.66-2.07) | 1.18 (0.62-2.23) |
|  | Diverse | 3 (100.0) | 0 (0.0) | undefined | undefined |
| **Age (years)** | 2-5 | 26 (89.7) | 3 (10.3) | 1.0 (ref) | 1.0 (ref) |
|  | 6-11 | 80 (87.9) | 11 (12.1) | 1.18 (0.35-3.94) | 1.27 (0.36-3.96) |
|  | 12-17 | 62 (71.3) | 25 (28.7) | 2.84 (0.92-8.72) | 2.84 (0.85-9.43) |
| **Parental education** | Tertiary | 99 (88.4) | 13 (11.6) | 1.0 (ref) | 1.0 (ref) |
|  | Secondary | 58 (74.4) | 20 (25.6) | 2.10 (1.11-3.98)*<sup>c</sup> | 2.10 (1.07-4.12)*<sup>c</sup> |
|  | Primary | 11 (64.7) | 6 (35.3) | 2.98 (1.31-6.79)*<sup>c</sup> | 2.97 (1.11-7.96)*<sup>c</sup> |
| **Financial situation** | High | 114 (85.7) | 19 (14.3) | 1.0 (ref) | 1.0 (ref) |
|  | Average to poor | 39 (72.2) | 15 (27.8) | 2.19 (1.22-3.09)*<sup>c</sup> | 2.18 (1.09-4.35)*<sup>c</sup> |
|  | Declined to answer | 15 (75.0) | 5 (25.0) | 1.54 (0.67-3.53)<sup>c</sup> | 1.55 (0.68-3.52)<sup>c</sup> |

<sup>a</sup>Prevalence ratio and 95% confidence interval are from Poisson regression with robust variance, and are adjusted for age, sex, or both according to independent variable.
<sup>b</sup>Prevalence ratio and 95% confidence interval are from Poisson regression with random effect on the household using the GLMMadaptive package in R and are adjusted for age, sex, or both according to independent variable.
<sup>c</sup>Adjusting for age and sex
\* indicates p-value <0.05
\*\*indicates p-value <0.01

## Discussion

In this population-based study, 13.3% of children experienced symptoms lasting more than 2 weeks since the beginning of the COVID-19 pandemic. Stratifying by age groups, the proportion of children experiencing persistent symptoms was significantly different (20.1 percentage points higher) between seropositive and seronegative adolescents (12 to 17 years old). For symptoms lasting more than 4 weeks among this age group, the adjusted prevalence difference between seronegatives and seropositives was 4.4%. No difference in reported persistent symptoms was observed between the seropositive and seronegative children younger than 12 years old.

In our study, we investigated symptoms that lasted at least 2 weeks, and we assessed a large variety of potential post-SARS-CoV-2 infection symptoms. Among children, there is currently no generalized definition, nor a diagnostic test of long COVID, and no precise understanding of the duration of symptoms that justifies the diagnosis. According to the World Health Organization, long COVID is considered when persistent symptoms last at least 12 weeks in adults but this definition may be different for children [2]. Among children, symptoms lasting up to 4 weeks are considered as potentially acute or subacute symptoms, and most studies on long COVID among children have used a duration ranging from 4 to 8 weeks to justify the diagnosis. In our study, the majority of children declared symptoms lasting 2 to 4 weeks, suggesting that symptoms were mostly acute or subacute and not long-lasting.

Cross-sectional studies on long COVID among children[3], [12-14] have reported prevalence estimates of persistent symptoms lasting more than 4 weeks ranging from 4.6% to 24.0%, with a strong heterogeneity in study design, inclusion criteria and outcomes. In our sample, 3.0% of seropositive children were declared as having symptoms lasting longer than 4 weeks; this proportion was 5.5% among seropositive adolescents aged 12 to 17, and 1.1% among seronegative participants in the same age group. This suggests that the prevalence of long COVID, defined as persistent symptoms lasting more than 4 weeks, may be around 4.4% in adolescents aged 12 to 17. Long-COVID as per this definition seems to affect fewer children below the age of 12 years in our sample. However, it needs to be noted that our sample does not allow for enough statistical power to capture small proportions; for this a much larger sample would be needed.

Importantly, our analysis is based on a population-based sample where serological status was used to identify previous infection. Compared to clinical studies or studies of PCR-confirmed cases, this design has the advantage of including severe, mild and asymptomatic infections, thereby yielding a more accurate denominator of the proportion of children infected. The latter may also explain why our estimated prevalence of long COVID is low compared to other studies. It also gives a seronegative control group, which allows us to distinguish symptoms that may be due to long COVID from symptoms due to other viruses or to the pandemic context (school closures, fewer social interactions, being unable to do sports and other activities or seeing family and friends suffering from COVID-19)[21].

Our results highlight the importance of stratifying by age groups when examining long COVID in children, as the prevalence and characteristics of long COVID likely vary between younger children and adolescents. Recent studies show that one third of adults present symptoms 30 to 45 days after their infection and those symptoms can persist for many months [22]. Our estimation of 4.4% prevalence of symptoms lasting over 4 weeks in the 12 to 17 age group suggests that adolescents remain less likely than adults to experience long-term symptoms, although this number still represents a large number of adolescents in absolute terms. The overall duration of long COVID in children and adolescents still needs to be studied and defined.

The lives of children and adolescents have been strongly disrupted since the beginning of the pandemic and concerns have been raised about the long term mental and physical health impact of the crisis [23]. In addition, evidence of persistent symptoms among children and a relatively large proportion declaring having had severe symptoms that resulted in a daily burden (even if not all long term), highlight the importance of monitoring the physical and mental impact of COVID-19 on this generation of children and adolescents.

In analyses restricted to seropositive children, we highlighted that children from households with a disadvantaged socioeconomic background were more likely to report symptoms lasting at least 2 weeks. To our knowledge, no previous study has assessed socioeconomic inequalities in long COVID in children nor in adults. These findings are not surprising as they reflect an extensive body of literature linking socioeconomic conditions to several negative health outcomes in children and adolescents [24].

The major strength of this study is that it relies on a randomly selected population-based sample. To date, very few studies on long COVID among children are population-based, and include children from the age of 2. Previous SARS-CoV-2 infection was assessed with an objective measure, enabling us to benefit from a seronegative control group. Relying on serological tests rather than only on confirmed infections gives the advantage of including mild and asymptomatic COVID-19 cases and to get a more precise denominator for SARS-CoV-2 infections. Also, it prevents the bias of participants over-reporting persistent symptoms when knowing they have been infected [25]. To our knowledge, only one previous study on long COVID in children was based on a serosurvey [14].

The study also has several limitations. First, it relies on parent-reported data on questionnaires without clinical assessment and objective measures. The presence and duration of persistent symptoms among children is based on the parental point of view, which can be subjective due to a potential memory bias and influenced by the parent’s background as well as the general household environment [21]. Also, the lack of information on the duration of persistent symptoms after four weeks as well as symptoms’ daily burden not being based on a standardized measure complicates the classification and diagnosis of long COVID. Finally, our sample is relatively small, with less than 10% of power to detect a difference of 1% in the prevalence of symptoms lasting more than 4 weeks between the seropositive and the seronegative sample.

## Conclusion

Our findings revealed that a significant proportion of children aged 12 to 17 years have symptoms lasting over two weeks after SARS-COV-2 infection, as assessed by serological status before vaccination. The estimated prevalence of symptoms lasting over 4 weeks is of 4.4% in this age group which suggests that adolescents are less likely than adults to experience long-term symptoms. This proportion represents a large number of adolescents in absolute terms, and should raise concern in the context of unknown long term evolution of symptoms. Children aged 2 to 11 years appear to experience fewer long-lasting symptoms related to SARS-CoV-2 infection, as no difference was observed between the seropositive and seronegative sample. However, our power to detect small differences was limited by the sample size, and further larger studies are needed to assess the prevalence of persistent symptoms among younger children. Monitoring the evolution of long COVID among children and adolescents is highly important as the long term physical and mental impact of COVID-19 persistent symptoms remains unclear and adequate public health policies are needed in terms of schooling and vaccination.

## Data Availability

Our data are accessible to researchers upon reasonable request for data sharing to the corresponding author.

## Acknowledgments

We are grateful to the staff of the Unit of Population Epidemiology of the HUG Division of Primary Care Medicine as well as to all participants whose contributions were invaluable to the study.

## List of abbreviations

ΔaPrev: adjusted Prevalence Difference
aPrev: adjusted Prevalence
aPR: adjusted Prevalence Ratio
CI: Confidence interval
COVID-19: Coronavirus disease 2019
SARS-CoV-2: Severe acute respiratory syndrome coronavirus 2

## Notes

**Conflict of interest** None of the funding institutions participated in the conduct of the research or in the preparation of the article.

### Competing Interest Statement

The authors have declared no competing interest.

### Funding Statement

Swiss Federal Office of Public Health, Geneva General Directorate of Health, HUG Private Foundation, Swiss School of Public Health, Fondation des Grangettes.

### Author Declarations

The Specchio-COVID19 study was approved by the Cantonal Research Ethics Commission of Geneva, Switzerland (CCER Project ID 2020-00881).

