## Supplemental Table 1 for "Persistent symptoms among children and adolescents with and without anti-SARS-CoV-2 antibodies: a population-based serological study in Geneva, Switzerland"

Isabelle Arm-Vernez, Andrew S Azman, Fatim Ba, Oumar Ba, Delphine Bachmann, Antoine Bal, Jean-François Balavoine, Michael Balavoine, Rémy Barbe, Hélène Baysson, Lison Beigbeder, Julie Berthelot, Patrick Bleich, Gaëlle Bryand, François Chappuis, Prune Collombet, Delphine Courvoisier, Alain Cudet, Carlos de Mestral Vargas, Paola D'ippolito, Richard Dubos, Roxane Dumont, Isabella Eckerle, Nacira El Merjani, Antoine Flahault, Natalie Francioli, Marion Frangville, Idris Guessous, Séverine Harnal, Samia Hurst, Laurent Kaiser, Omar Kherad, Julien Lamour, Pierre Lescuyer, François L'Huissier, Fanny-Blanche Lombard, Andrea Jutta Loizeau, Elsa Lorthe, Chantal Martinez, Lucie Ménard, Lakshmi Menon, Ludovic Metral-Boffod, Benjamin Meyer, Alexandre Moulin, Mayssam Nehme, Natacha Noël, Francesco Pennacchio, Javier Perez-Saez, Didier Pittet, Jane Portier, Klara M Posfay-Barbe, Géraldine Poulain, Caroline Pugin, Nick Pullen, Zo Francia Randrianandrasana, Aude Richard, Viviane Richard, Frederic Rinaldi, Jessica Rizzo, Khadija Samir, Claire Semaani, Silvia Stringhini, Stéphanie Testini, Jennifer Villers, Guillemette Violot, Nicolas Vuilleumier, Ania Wisniak, Sabine Yerly, María-Eugenia Zaballa

**Supplementary Figure S1**: Study flow diagram.

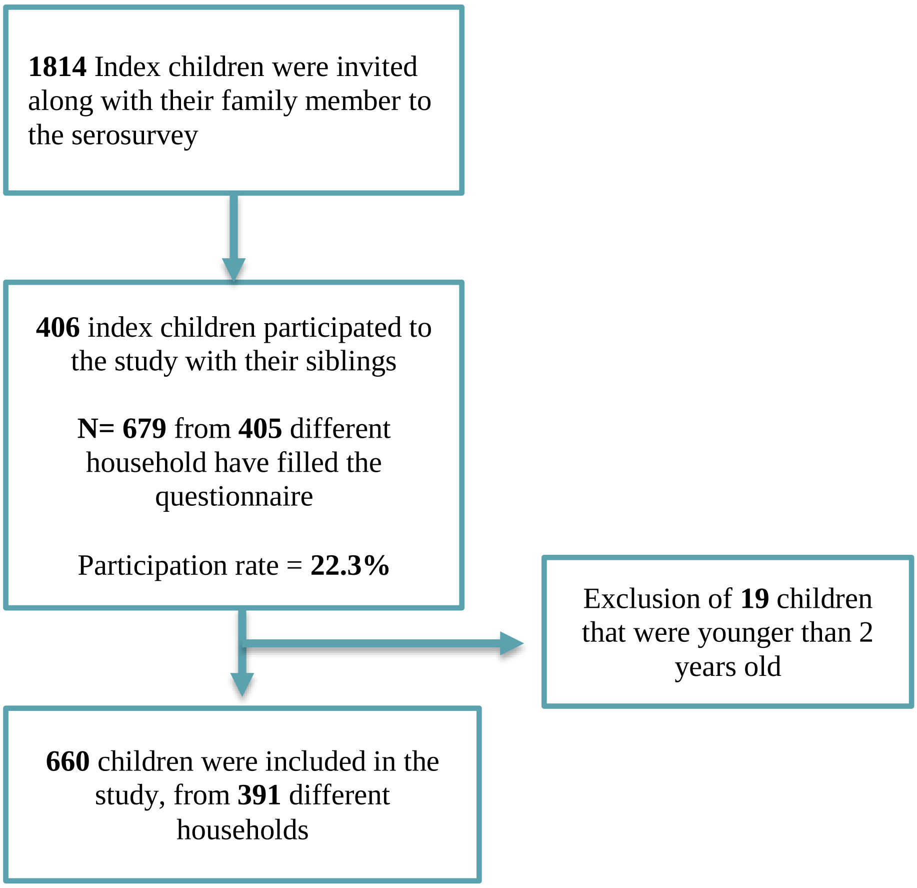

**Supplementary Figure S2: Age- and sex-adjusted prevalence of persistent symptoms, stratified by age group and serological status**

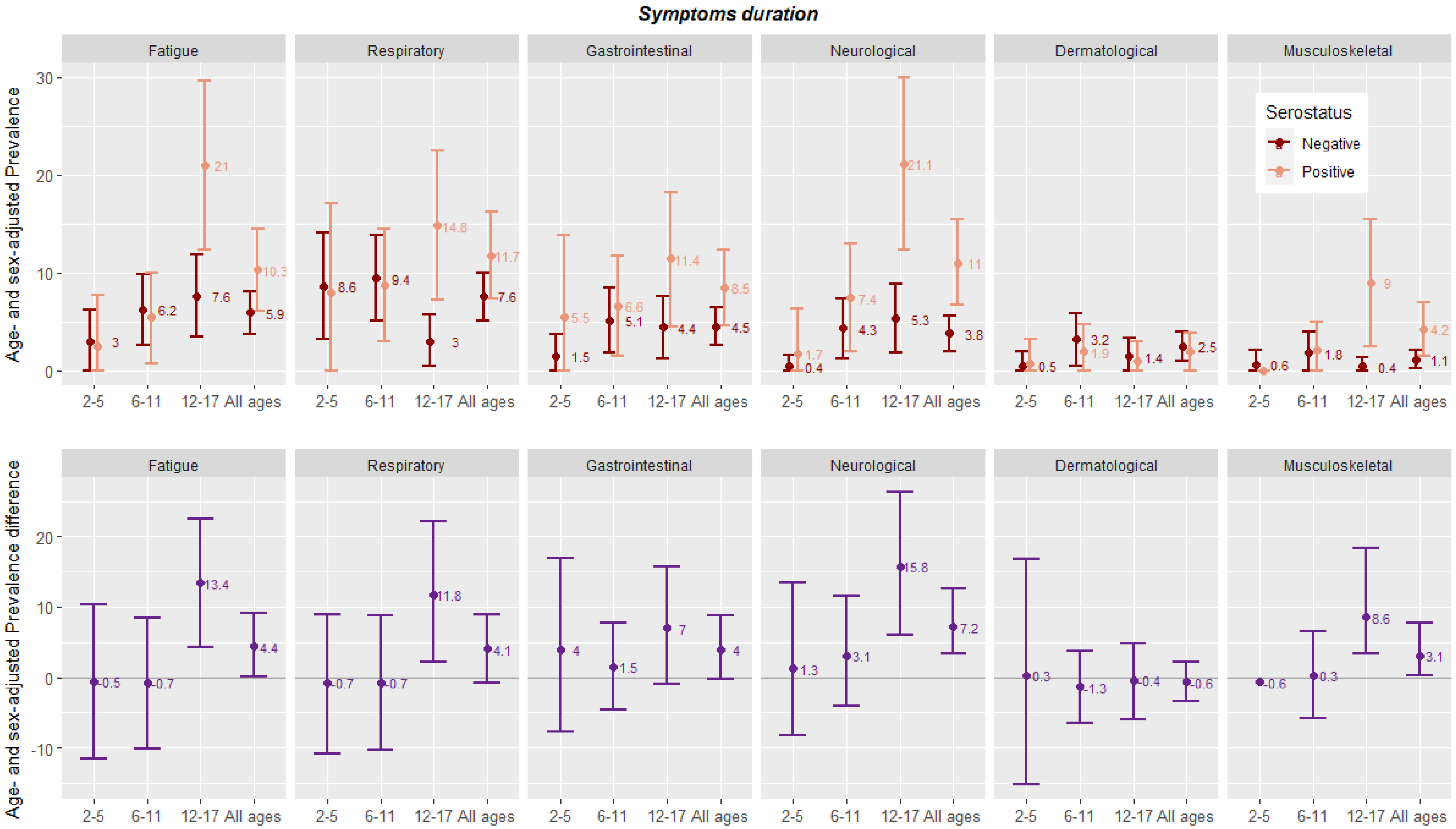

### Table S1. Description of symptoms

| Symptoms[a] | **Seronegative** n (%) | **Seropositive** n (%) | p-value[b] |
|---|---|---|---|
| **N** | 453 | 207 | |
| Fatigue | 28 (6.2) | 24 (11.5) | **0.026** |
| ***Respiratory symptoms:*** | | | |
| Cough | 19 (4.2) | 11 (5.3) | 0.665 |
| Sore throat | 14 (3.1) | 11 (5.3) | 0.245 |
| Runny nose | 27 (5.9) | 13 (6.2) | 1.000 |
| Difficulty breathing | 1 (0.2) | 2 (1.0) | 0.487 |
| Fever | 13 (2.9) | 13 (6.2) | 0.062 |
| ***Gastrointestinal symptoms*** | | | |
| Loss of appetite | 11 (2.4) | 7 (3.4) | 0.664 |
| Nausea | 5 (1.1) | 5 (2.4) | 0.351 |
| Vomiting | 2 (0.4) | 1 (0.5) | 1.000 |
| Constipation | 4 (0.9) | 4 (1.9) | 0.450 |
| Diarrhea | 5 (1.1) | 6 (2.9) | 0.181 |
| Weight loss | 1 (0.2) | 1 (0.5) | 1.000 |
| Stomach pain | 10 (2.2) | 6 (2.9) | 0.797 |
| ***Musculoskeletal symptoms*** | | | |
| Muscle pain | 7 (1.5) | 10 (4.8) | 0.028 |
| Joint pain | 2 (0.4) | 3 (1.4) | 0.369 |
| ***Neurological Symptoms*** | | | |
| Headache | 13 (2.9) | 23 (11.1) | **<0.001** |
| Loss of smell | 0 (0.0) | 10 (4.8) | **<0.001** |
| Loss of taste | 0 (0.0) | 8 (3.8) | **<0.001** |
| Dizziness | 0 (0.0) | 1 (0.5) | 0.689 |
| Changes in vision | 1 (0.2) | 0 (0.0) | 1.000 |
| Shaking | 1 (0.2) | 1 (0.5) | 1.000 |
| Tingling | 1 (0.2) | 0 (0.0) | 1.000 |
| Confusion | 4 (0.9) | 4 (1.9) | 0.450 |
| Insomnia | 6 (1.3) | 2 (1.0) | 0.992 |
| Hypersomnia | 0 (0.0) | 2 (1.0) | 0.184 |
| ***Cardiovascular symptoms*** | | | |
| Palpitations | 0 (0.0) | 1 (0.5) | 0.689 |
| ***Dermatological symptoms*** | | | |
| Skin rashes | 4 (0.9) | 4 (1.9) | 0.450 |
| Itching skin | 5 (1.1) | 2 (1.0) | 1.000 |
| Redness skin | 5 (1.1) | 1 (0.5) | 0.734 |

[a]**Answers based on the question: "**Since the start of the pandemic in Switzerland, has the child developed one or more symptoms (including fatigue) that have lasted at least two weeks?" If yes, which ones?

[b] Fisher's exact and Pearson's Chi-squared test between seronegative and seropositive

### Table S2. Age- and sex-adjusted prevalence of persistent symptoms, stratified by age group and serological status

| | 2-5 years old | | | 6-11 years old | | | 12-17 years old | | |
|---|---|---|---|---|---|---|---|---|---|
| | Seronegative N=118 | Seropositive N=29 | | Seronegative N=180 | Seropositive N=91 | | Seronegative N=155 | Seropositive N=87 | |
| | aPrev[a,b] | aPrev[a,b] | ΔaPrev | aPrev[a,b] | aPrev[a,b] | ΔaPrev | aPrev[a,b] | aPrev[a,b] | ΔaPrev |
| **Symptoms > 2 weeks** | 8.6 (3.2,14.1) | 7.9 (0.0,17.1) | -0.7 (-13.4,11.6) | 12.4 (7.5,17.3) | 11.9 (5.2,18.6) | -0.5 (-8.2,7.1) | 8.9 (4.4,13.4) | 29.0 (19.4,38.7) | 20.1** (10.6,29.7) |
| **Reported symptoms** | | | | | | | | | |
| Fatigue | 3.0 (0.0,6.2) | 2.5 (0.0,7.7) | -0.5 (-11.6,10.4) | 6.2 (2.6,9.8) | 5.5 (0.7,10) | -0.7 (-8.7,7.1) | 7.6 (3.4,11.8) | 21.0 (12.3,29.6) | 13.4** (4.7,22.1) |
| Respiratory | 8.6 (3.2,14.1) | 7.8 (0.0,17.1) | -0.7 (-16.2,14.5) | 9.4 (5.1,13.8) | 8.7 (2.9,14.5) | -0.7 (-8.3,6.8) | 3 (0.4,5.7) | 14.8 (7.2,22.5) | 11.8* (2.7,21.6) |
| Gastrointestinal | 1.5 (0.0,3.7) | 5.5 (0.0,13.8) | 4.0 (-8.3,17.5) | 5.1 (1.8,8.5) | 6.6 (1.4,11.7) | 1.5 (-4.8,8.2) | 4.4 (1.2,7.6) | 11.4 (4.5,18.2) | 7.0 (-0.8,15.7) |
| Neurological | 0.4 (0.0,1.6) | 1.7 (0.0,6.3) | 1.3 (-9.3,14.5) | 4.3 (1.2,7.3) | 7.4 (1.9,12.9) | 3.1 (-4.7,12.2) | 5.3 (1.8,8.8) | 21.1 (12.3,29.9) | 15.8** (5.6,26.7) |
| Dermatological | 0.5 (0.0,2.0) | 0.8 (0.0,3.2) | 0.3 (-8.0,9.5) | 3.2 (0.5,5.8) | 1.9 (0.0,4.7) | -1.3 (-6.8,3.9) | 1.4 (0.0,3.3) | 1.0 (0.0,3.0) | -0.4 (-5.6,4.5) |
| Musculoskeletal | 0.6 (0.0,2.1) | 0.0 (0,0) | -0.6 undefined | 1.8 (0.0,4.0) | 2.1 (0.0,4.9) | 0.3 (-4.9,5.5) | 0.4 (0.0,1.3) | 9.0 (2.4,15.5) | 8.6** (3.9,18) |
| **Symptoms duration** | | | | | | | | | |
| 2 to 3 weeks | 2.7 (0.0,5.7) | 7.1 (0.0,16.4) | 4.4 (-6.3,17.2) | 8.2 (4.1,12.2) | 17.7 (9.8,25.6) | 9.5* (1.9,17.3) | 3.0 (0.4,5.7) | 24.4. (15.2,33.6) | 21.4** (12.0,31.4) |
| 3 to 4 weeks | 0.5 (0.0,1.9) | 0.0 (0.0,0.0) | -0.5 undefined | 1.4 (0.0,3.2) | 0.0 (0.0,0.0) | -1.4 undefined | 0.1 (0.0,0.5) | 0.4 (0.0,1.2) | 0.3 (1.5,16.6) |
| More than 4 weeks | 3.8 (0.2,7.5) | 0.0 (0.0,0.0) | -3.8 undefined | 4.4 (1.4,7.4) | 0.0 (0.0,0.0) | -4.4 undefined | 1.1 (0.0,2.6) | 5.5 (0.5,10.3) | 4.4 (-3.8,13.6) |

[a] Prevalence was computed using marginal prediction of logistic regression and 95% CI were computed using normal approximation and truncated to 0 for the aPRev.
[b] Prevalence was adjusted for sex and age
* indicates p-value <0.05
**indicates p-value <0.01

### Additional information on the serological test

The serological tests were based on commercially-available immunoassays Roche Elecsys anti-SARS-CoV-2 N immunoassay, which detects immunoglobulins (IgG/A/M) targeting the virus nucleocapsid (N) protein (#09 203 079 190, Roche-N) and has an in-house sensitivity of 99.8% (95% CrI, 99.4%-100%) and specificity of 99.1% (95% CI, 98.3%-99.7%) (Roche Diagnostics, Rotkreuz, Switzerland).We defined seropositivity using the manufacturer's provided cut-off index ≥1.0 for the Roche-N immunoassays.
